# Assessing the impact of community-based interventions on hypertension and diabetes management in three Minnesota communities: findings from the prospective evaluation of US HealthRise programs

**DOI:** 10.1101/2021.08.26.21262688

**Authors:** Nancy Fullman, Krycia Cowling, Luisa S Flor, Shelley Wilson, Paurvi Bhatt, Miranda F Bryant, Joseph N Camarda, Danny V Colombara, Jessica Daly, Rose K Gabert, Katie Panhorst Harris, Casey K Johanns, Charlie Mandile, Susan Marshall, Claire R McNellan, Vasudha Mulakaluri, Bryan K Phillips, Marissa B Reitsma, Naomi Sadighi, Tsega Tamene, Blake Thomson, Alexandra Wollum, Emmanuela Gakidou

**Author notes:** Corresponding author: Nancy Fullman, Institute for Health Metrics and Evaluation, Department of Health Metrics Sciences, University of Washington, Seattle, WA 98195, USA.

## Abstract

**Background:** Community-based health interventions are increasingly viewed as models of care that can bridge healthcare gaps experienced by underserved communities in the United States (US). With this study, we sought to assess the impact of such interventions, as implemented through the US HealthRise program, on hypertension and diabetes among underserved communities in Hennepin, Ramsey, and Rice Counties, Minnesota.

**Methods and findings:** HealthRise patient data from June 2016 to October 2018 were assessed relative to comparison patients in a difference-in-difference analysis, quantifying program impact on reducing systolic blood pressure (SBP) and hemoglobin A1c, as well as meeting clinical targets (< 140 mmHg for hypertension, < 8% Al1c for diabetes), beyond routine care. For hypertension, HealthRise participation was associated with SBP reductions in Rice (6.9 mmHg [95% confidence interval: 0.9–12.9]) and higher clinical target achievement in Hennepin (27.3 percentage-points [9.8–44.9]) and Rice (17.1 percentage-points [0.9 to 33.3]). For diabetes, HealthRise was associated with A1c decreases in Ramsey (1.3 [0.4–2.2]). Qualitative data showed the value of home visits alongside clinic-based services; however, challenges remained, including community health worker retention and program sustainability.

**Conclusions:** HealthRise participation had positive effects on improving hypertension and diabetes outcomes at some sites. While community-based health programs can help bridge healthcare gaps, they alone cannot fully address structural inequalities experienced by many underserved communities.

## Introduction

Longstanding health disparities occur throughout the US,[1] with differences often manifesting across multiple dimensions (e.g., geography, gender, race or ethnicity, socioeconomic status).[2] Non-communicable diseases (NCDs) and NCD-related risks like hypertension can uniquely affect underserved communities, as a constellation of factors – from service affordability and inadequate insurance to more entrenched socioeconomic obstacles like low access to nutritional food – can quickly give way to high rates of chronic, debilitating conditions without good access to care or appropriate services. Community-based interventions with integrated care have emerged as approaches to bridge gaps in NCD care, particularly for underserved areas in the US.[3–9] Nonetheless, the effectiveness of these interventions vary across settings; interventions implemented; roles of community health workers (CHWs)(e.g., direct involvement in patient care[10] *versus* assisting health providers who are then responsible for service provision[11]); and NCDs targeted. While CHWs and community-based interventions appear to be promising models of care for hypertension and diabetes in underserved communities,[5–7] more rigorous evaluations across more local contexts and populations are needed.

To strengthen this evidence base, the HealthRise program was developed to implement and pilot locally-tailored programs for improving screening, diagnosis, management and control of hypertension and diabetes among underserved communities.[12–14] Funded by the Medtronic Foundation, HealthRise took place in nine communities in four countries – Brazil, India, South Africa, and the US – from 2014 to 2018. In the US, Minnesota was selected, a state which generally surpasses national averages and ranks among the healthiest across many health measures.[1,15] Nonetheless, county- and sub-county level health disparities remain in Minnesota, particularly among populations that face compounding barriers to care and improved health outcomes.[15,16] To maximize the potential impact of HealthRise programs, especially within a relatively short time span (i.e., the earliest US program began in 2016), HealthRise targeted geographic areas with the greatest need and highest disease burden. A 2014-2015 needs assessment identified communities in three counties – Hennepin, Ramsey, and Rice – as candidates for HealthRise programs based on their high NCD burden and risk profiles, challenges in healthcare access, and sociodemographic characteristics (e.g., large minority and/or immigrant populations).[15,17] Three main recommendations emerged from this assessment: (1) focus on people with the highest need and poorest clinical outcomes (2) use multi-faceted interventions to address multiple risks and comorbid conditions; and (3) identify opportunities to integrate CHWs within formal health system functions. Based on this needs assessment and grantee applications, three implementing partners were selected (i.e., one per county), community-based interventions were developed, and US HealthRise program implementation took place from June 2016 to October 2018.

With this study, we provide key findings from HealthRise programs in Hennepin, Ramsey, and Rice Counties, Minnesota. Based on quantitative and qualitative data collected over the course of program implementation, we evaluated the potential impact of these community-based interventions on improving clinical and health outcomes for hypertension and diabetes patients. We conducted difference-in-difference analyses in relation to comparison patients to quantify this impact above and beyond what might be expected for demographically similar patients under routine care in the same communities. This study contributes to the science supporting the role of community-based programs in elevating the health of underserved communities, in the US and elsewhere.

## Materials and Methods

### Study overview, design, and interventions

This analysis follows the global HealthRise prospective evaluation framework, which was established in 2014 and agreed upon by all partners; greater detail on the global team structure, interventions, and analyses are provided elsewhere.[12–14] In sum, HealthRise was funded by the Medtronic Foundation, with global and US implementation partners coordinated by Abt Associates and evaluation activities overseen by the Institute for Health Metrics and Evaluation (IHME). Though ongoing coordination and collaboration occurred across implementation and evaluation organizations, these grant streams were purposefully structured and funded separately to support an independent assessment of the HealthRise programs.

For US HealthRise, patient-level monitoring data were routinely collected and collated by grantees during program implementation (June 2016 to October 2018). Using a mixed-methods quasi-experimental design, we synthesized qualitative and quantitative data from HealthRise and comparison patients and stakeholders (e.g., service providers, administrators, and policymakers) to inform its endline evaluation. Table 1 summarizes key information on each US HealthRise site and interventions implemented by grantee, as interventions were tailored to address gaps or challenges identified for each site during the 2014-2015 needs assessment;[15,17] more detail is available elsewhere.[14]

**Table 1.**
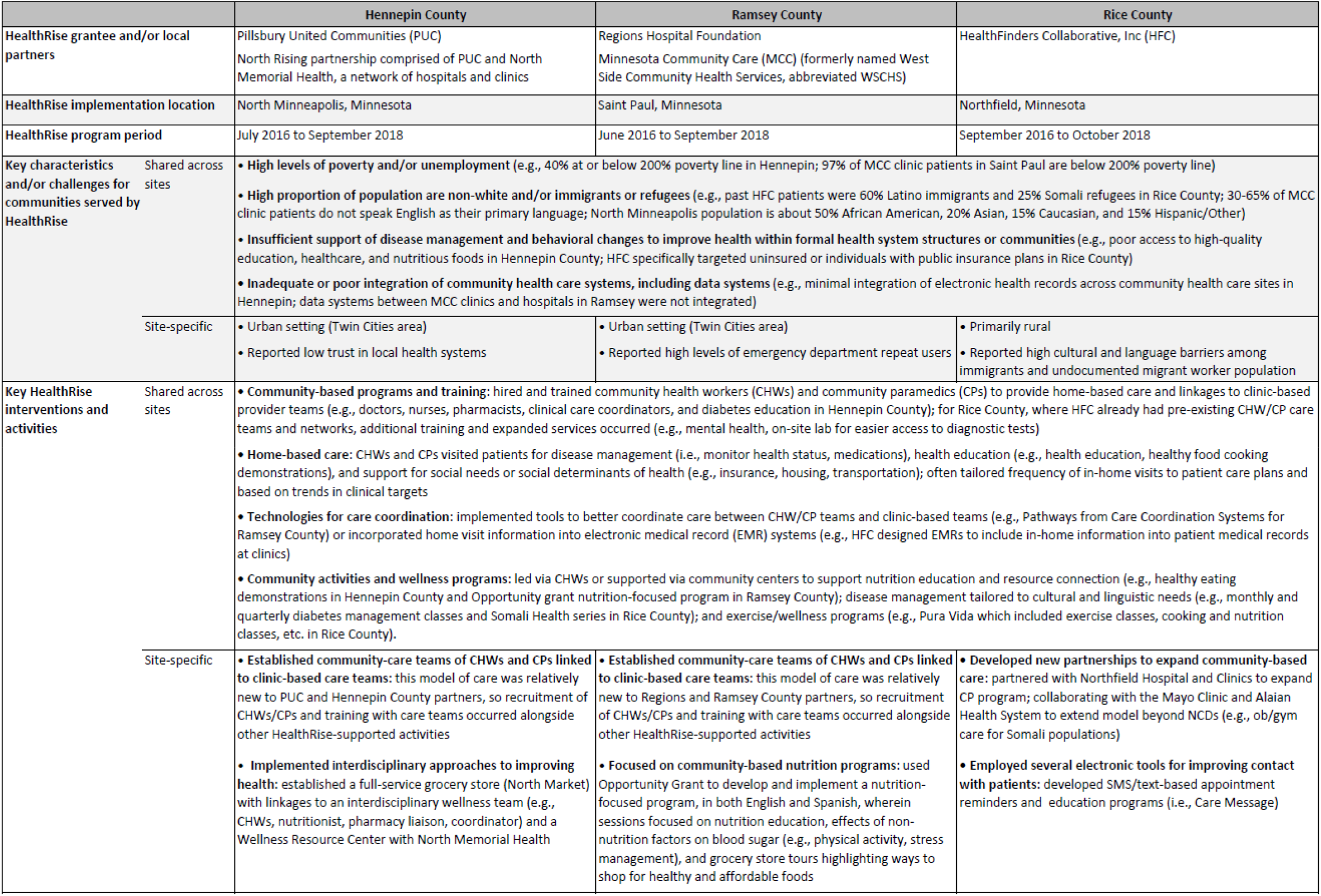
Overview of interventions by US HealthRise site. More detailed descriptions of HealthRise interventions, as provided by grantees and compiled by Abt Associates, are available elsewhere.[14]

### Definitions and data

#### Definitions

We used following case definitions for hypertension and diabetes at each time point: (1) prevalent cases were patients with documented diagnoses, or patients without prior diagnoses but with clinical readings that would qualify for diagnosis (i.e., systolic blood pressure [SBP] ≥ 140 mmHg or diastolic blood pressure [DBP] ≥ 90 mmHg for hypertension; hemoglobin A1c ≥ 6.5% for diabetes); (2) diagnosed cases were patients with documented diagnoses; and (3) patients meeting treatment targets were prevalent cases with SBP < 140 mmHg and DBP < 90 mmHg for hypertension, and A1c < 8% for diabetes. If DBP measures were not available for a given patient, then only SBP readings were used.

#### Endline evaluation data collection

##### HealthRise patient data

Each US grantee collected patient-level data from existing sources and provided de-identified data over time. To best capture potential program impact, analyses were limited to HealthRise patients who (1) remained enrolled in HealthRise at endline (i.e., never withdrew from programs); and (2) had at least two separate biometric data points for blood pressure (i.e., ideally both SBP and DBP, but at minimum, SBP) or A1c (Table 2). Subsequently, evaluation results reflected potential effects from HealthRise participation, and not “intention to treat,” which would have included patients who enrolled but then withdrew from the program at some point. Rates of any program withdrawal varied by site, ranging from 16.7% (n=19) for Hennepin to 32.5% (n=25) for Ramsey.

**Table 2.**
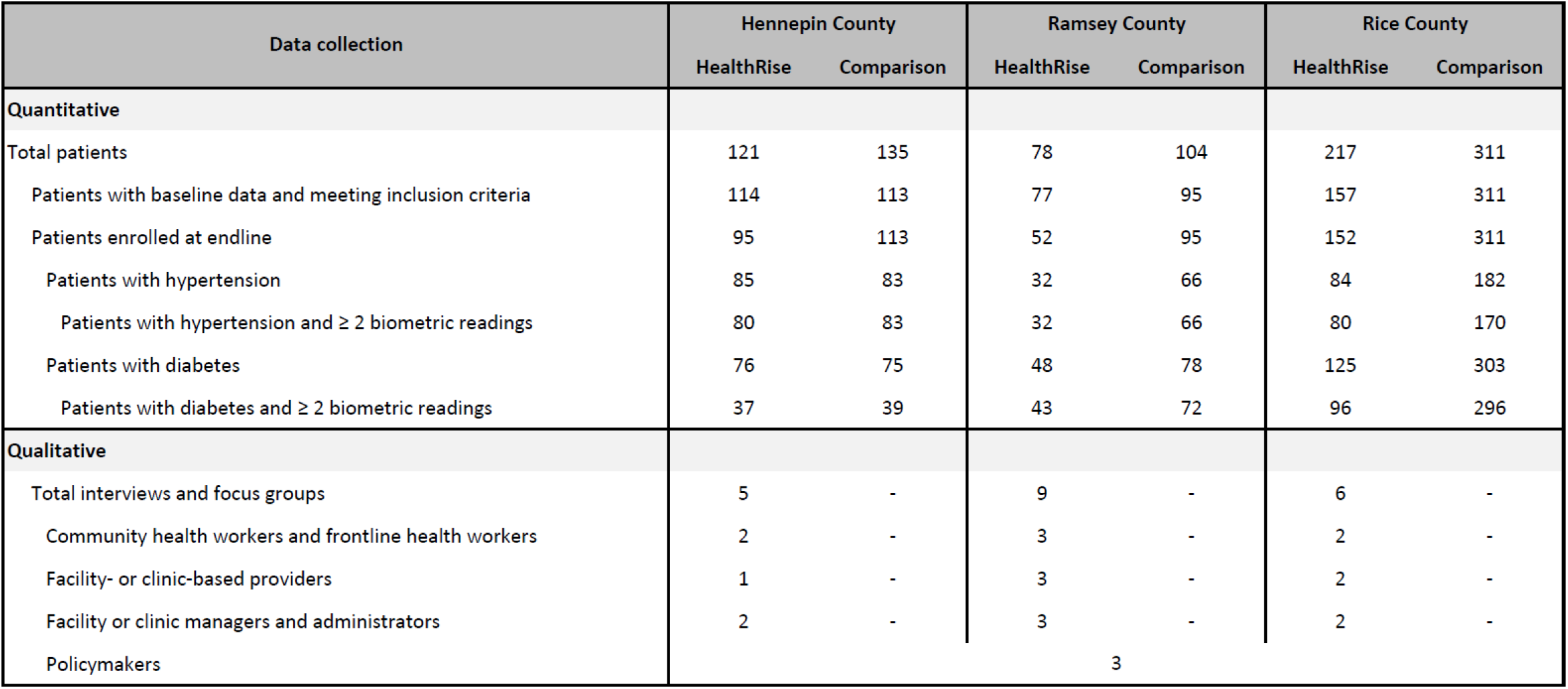
Endline data availability and patient sample sizes by HealthRise site and for intervention and comparison patients. “Total patients” reflect the number of patients included in original samples, irrespective of additional inclusion or exclusion criteria. “Patients with baseline data and meeting inclusion criteria” reflect the number of patients who met inclusion criteria at baseline (i.e, prevalent case of hypertension or diabetes and aged 30-89 years old); for HealthRise patients, this also reflects the number of patients who formally enrolled in the program, whereas comparison patients were limited to individuals who could have met eligibility requirements at each site within the program implementation time periods (2016-2018). “Total patients enrolled at endline” reflect the number of patients who did not withdraw from HealthRise during program duration. At each site and group, some proportion of patients have both hypertension and diabetes.

For Rice, 3.2% of patients (n=5) lacked a clinic visit or biometric data since baseline, and thus were considered withdrawn. In Ramsey, most patients who withdrew did so after a few months of enrollment and within the first year of HealthRise implementation.

For baseline measures, we used biometric data collected at HealthRise program enrollment. If such data were not available at the precise enrollment date, then biometric data were used from the data closest to that of enrollment. For endline measures, we used patients’ most recent biometric measurements.

##### Comparison patient data

Grantees provided comparison data drawn from patient populations similar to those enrolled in HealthRise. Upon receiving each site’s dataset, we sought to reconstruct samples of comparison patients that were similar demographically and in terms of baseline health conditions to HealthRise patients (i.e., excluding comparison patients younger than 30 years and 90 years or older, and those without a diagnosis of hypertension or diabetes and had baseline biometric data that fell within disease control categories). As necessary, comparison patient data were censored to correspond with each site’s HealthRise program implementation period (Table 1), and thus better approximate similar follow-up times for comparison patients. After this censoring step, we then excluded any comparison patients who lacked more than one measurement of A1c and systolic blood pressure and therefore could not contribute to baseline *versus* endline comparisons. Included comparison patients, by site, are provided in Table 2.

Comparison patient data selection occurred between October 2018 and January 2019, with criteria determined by site. For Hennepin, data for patients who formed the comparison group were extracted from clinics associated with North Memorial but had not enrolled in HealthRise. Selection criteria included having at least two biometric readings for A1c or SBP – one in 2016 and one in 2018 – to approximate baseline and endline measures for HealthRise; and being between the ages of 30 and 89 years at “baseline.” For Ramsey, comparison patient data were extracted through Minnesota Community Care (formerly named West Side Community Health Services); eligible individuals were patients who had not enrolled in HealthRise and had similar baseline levels of A1c or SBP as HealthRise patients. For Rice, data were extracted from a partner clinic where HealthRise interventions were not implemented. Unlike other comparison patient datasets, International Classification of Disease codes for diabetes and hypertension were not available for patient diagnosis; instead, the diagnosis variable for Rice comparison patients listed active diagnoses. Consequently, a text-matching algorithm was applied to assign diabetes and/or hypertension diagnosis based on the text data in this variable.

##### Qualitative data

Twenty-three key informant interviews (KIIs) were conducted with local policymakers (non-site specific) and with different types of staff at each site (Table 2). Interviews were not conducted with patients or with staff at clinics from which comparison patient data were selected.

Initial potential interviewees were identified via leadership from HealthRise grantees and partner organizations, and then additional staff (e.g., clinic-based providers, community paramedics [CPs], CHWs) were contacted via snowball sampling. Of the original individuals identified, 79% completed one-hour interviews via telephone with an IHME evaluation team member.

All interviews were audio-recorded and listened to multiple times by a single researcher. Key components were transcribed in an Excel template, with thematic coding applied to identify both site-specific and overarching themes across sites.

#### Endline evaluation analyses

To quantify potential effects of HealthRise participation, we used two outcome indicators to measure patient-level changes from baseline to endline: (1) the proportion of patients meeting treatment targets (i.e., SBP < 140 mmHg and DBP < 90 mmHg for hypertension; < 8% A1c for diabetes); and (2) patient biometric measures (i.e., SBP for hypertension, A1c for diabetes). All analyses were limited to patients who were prevalent cases at baseline and had corresponding biometric data for each time point.

We conducted difference-in-difference analyses in two steps for each site and by condition. First, we ran an unadjusted model, only including binary variables for HealthRise status and timing (i.e., baseline or endline) and an interaction term for HealthRise at endline to capture the effect of HealthRise participation over time. We then ran an adjusted model, including the following covariates to account for potential systematic differences in HealthRise and comparison patients: sex (female, male); age (< 50 years, ≥ 50 years); time elapsed from baseline to endline (< 12 months, ≥ 12 months); and comorbidities at baseline (prevalent case of only hypertension or diabetes; prevalent case of both hypertension and diabetes). We specified robust standard errors for each model, and used Welch’s t-tests (i.e., assuming unequal variance between each group) to evaluate statistically significant differences between HealthRise and comparison patients. All analyses were conducted in Stata version 15 and R version 3.6.2.[18,19]

### Ethical approval

Ethical approval for this study was obtained from the University of Washington’s institutional review board, as well as the local data collection agencies and government entities for each site. All personal identifiers were removed prior to data sharing with IHME; only de-identified data were analyzed.

## Results

### Quantitative results

Overall, hypertension and diabetes indicators generally improved for HealthRise patients compared with baseline measures (Figure 1, Table 3). At the same time, clinical improvements were heterogeneous since program enrollment (Figure 1), emphasizing potential challenges in effective case management among underserved populations.

**Table 3.** Baseline and endline measures by HealthRise site and for intervention and comparison patients for hypertension (A) and diabetes (B). For HealthRise, samples reflect patients who were prevalent cases of hypertension or diabetes at baseline; remained enrolled throughout the program; and had at least two biometric readings during program participation.

**A) Hypertension**
| HealthRise program site | Sample size | Percent of patients meeting treatment targets (SBP < 140 and DBP < 90) (%) |  | Average SBP reading (mmHg) |  | Average changes in SBP from baseline to endline |  |
| --- | --- | --- | --- | --- | --- | --- | --- |
|  |  | Baseline | Endline | Baseline | Endline | Absolute change (mmHg) | Percent change (%) |
| Hennepin County |  |  |  |  |  |  |  |
| HealthRise patients | 80 | 53.8 (42.7 to 64.5) | 76.3 (65.6 to 84.4) | 136.2 (131.8 to 140.6) | 128.5 (125.6 to 131.5) | -7.7 (-12.1 to -3.2) | -4.2 (-7.2 to -1.1) |
| Comparison patients | 83 | 83.1 (73.3 to 89.8) | 78.3 (68.0 to 86.0) | 127.3 (124.3 to 130.3) | 125.3 (121.9 to 128.7) | -2.0 (-6.0 to 2.0) | -0.7 (-3.9 to 2.5) |
| Ramsey County |  |  |  |  |  |  |  |
| HealthRise patients | 32 | 12.5 (4.6 to 29.8) | 50.0 (32.7 to 67.3) | 150.4 (144.9 to 156.0) | 136.0 (126.6 to 145.5) | -14.4 (-23.0 to -5.8) | -9.4 (-15.0 to -3.9) |
| Comparison patients | 66 | 30.3 (20.3 to 42.6) | 45.5 (33.7 to 57.7) | 146.5 (141.5 to 151.6) | 142.9 (137.4 to 148.3) | -3.7 (-10.2 to 2.9) | -0.9 (-5.5 to 3.6) |
| Rice County |  |  |  |  |  |  |  |
| HealthRise patients | 80 | 42.5 (32.0 to 53.7) | 63.8 (52.5 to 73.6) | 142.5 (137.5 to 147.6) | 134.6 (129.9 to 139.3) | -7.9 (-13.4 to -2.5) | -4.0 (-7.7 to -0.4) |
| Comparison patients | 170 | 78.2 (71.4 to 83.8) | 82.4 (75.8 to 87.4) | 128.3 (126.0 to 130.6) | 127.3 (124.9 to 129.7) | -1.0 (-3.6 to 1.5) | 0.0 (-2.0 to 2.1) |

**B) Diabetes**
| HealthRise program site | Sample size | Percent of patients meeting treatment targets (A1c < 8%) |  | Average A1c reading (%) |  | Average changes in A1c from baseline to endline |  |
| --- | --- | --- | --- | --- | --- | --- | --- |
|  |  | Baseline | Endline | Baseline | Endline | Absolute change (%) | Percent change (%) |
| Hennepin County |  |  |  |  |  |  |  |
| HealthRise patients | 37 | 24.3 (12.9 to 41.1) | 24.3 (12.9 to 41.1) | 9.9 (9.1 to 10.7) | 9.4 (8.6 to 10.3) | -0.5 (-1.4 to 0.5) | -0.8 (-12.6 to 11.1) |
| Comparison patients | 39 | 71.8 (55.8 to 84.0) | 76.9 (60.7 to 87.8) | 7.6 (7.1 to 8.2) | 7.5 (6.9 to 8.1) | -0.1 (-0.7 to 0.5) | -0.2 (-6.3 to 5.9) |
| Ramsey County |  |  |  |  |  |  |  |
| HealthRise patients | 43 | 4.7 (1.1 to 17.4) | 25.6 (14.5 to 41.0) | 10.4 (9.7 to 11.2) | 9.0 (8.4 to 9.5) | -1.5 (-2.1 to -0.8) | -11.4 (-16.9 to -5.9) |
| Comparison patients | 72 | 26.4 (17.4 to 37.9) | 29.2 (19.7 to 40.8) | 10.0 (9.4 to 10.6) | 9.8 (9.2 to 10.4) | -0.2 (-0.8 to 0.5) | 2.7 (-4.9 to 10.3) |
| Rice County |  |  |  |  |  |  |  |
| HealthRise patients | 96 | 43.8 (34.1 to 53.9) | 51.0 (41.0 to 61.0) | 8.8 (8.3 to 9.2) | 8.5 (8.1 to 9.0) | -0.3 (-0.6 to 0.1) | -1.3 (-4.9 to 2.4) |
| Comparison patients | 296 | 50.3 (44.6 to 56.0) | 54.7 (49.0 to 60.3) | 8.6 (8.3 to 8.8) | 8.3 (8.1 to 8.6) | -0.2 (-0.5 to 0.0) | 0.1 (-2.4 to 2.6) |

**Figure 1.**
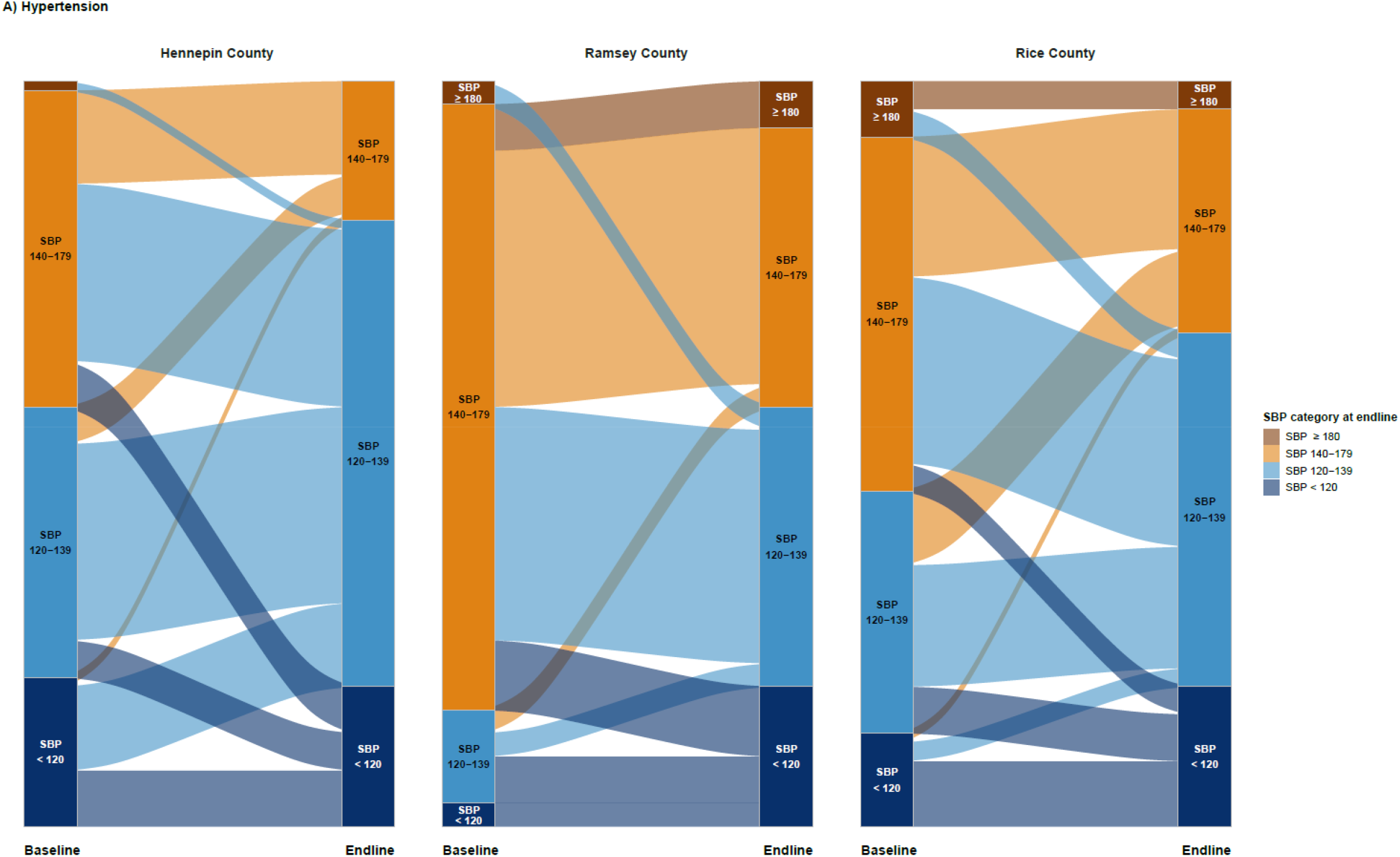
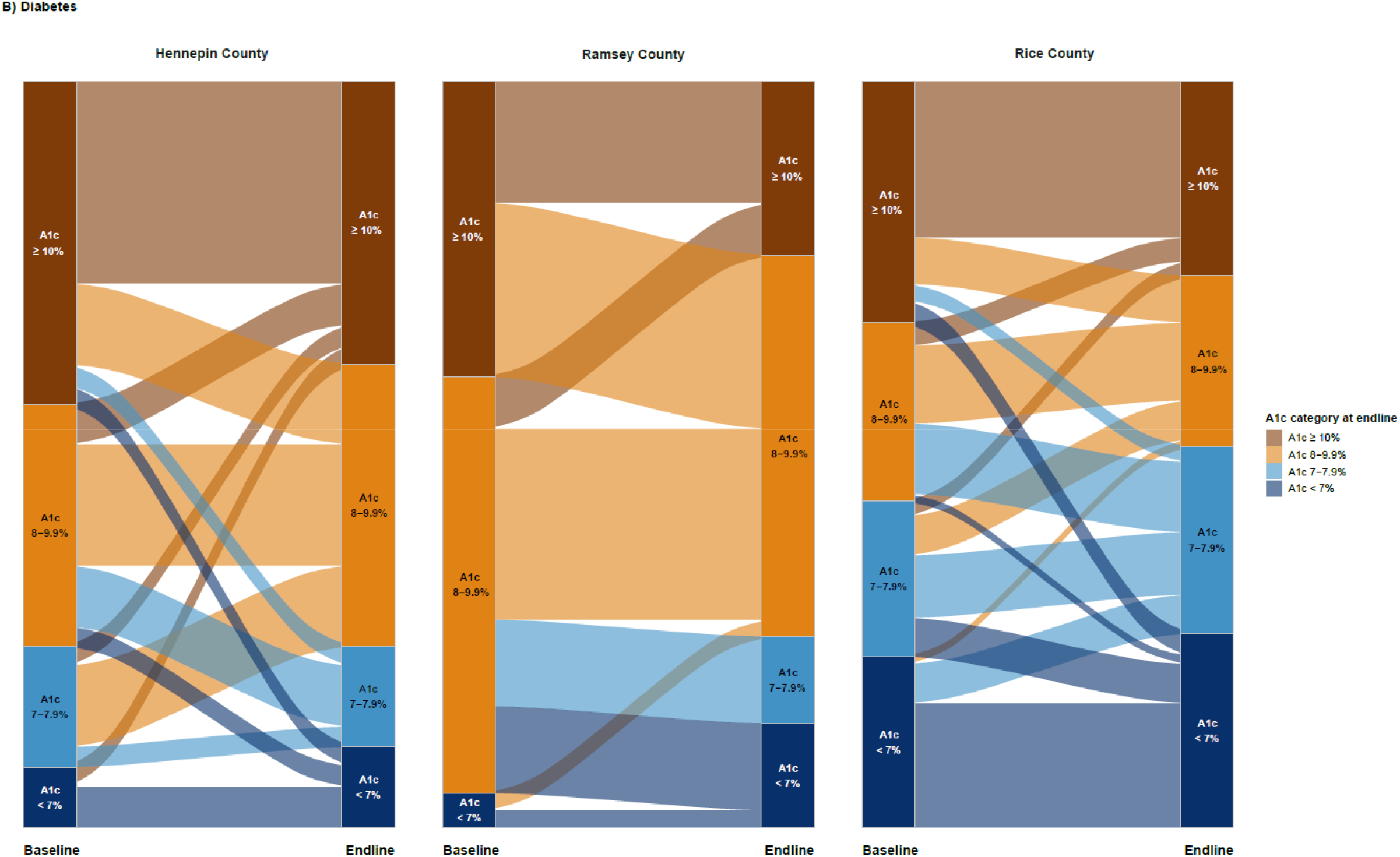
HealthRise patient shifts in disease severity categories between baseline and endline based on biometric readings for hypertension (A) and diabetes (B). The height of each column reflects 100% of patients at each time point (baseline and endline), while the categories within each column represents the percentage of patients in each category at baseline and endline. Patient groups are color-coded by their categorization at endline (right column per site) and flow from their categorization at baseline (left column per site)

Across sites, a considerable percentage of hypertension patients with baseline SBP measures exceeding 140 mmHg improved endline levels to below 140 mmHg (Figure 1A); this trend was particularly pronounced for Hennepin and Ramsey. In Hennepin, 76.8% (95% confidence interval: 65.6-84.4%) of hypertension patients enrolled in HealthRise recorded endline SBP measures below 140 mmHg and 50.0% (32.3-67.3%) of HealthRise patients with hypertension in Ramsey met this threshold. Nonetheless, some percentage of hypertension patients shifted into worse SBP categories by endline: 17.5% in Hennepin, 9.4% in Ramsey, and 13.8% in Rice. Sizeable improvements occurred for diabetes patients meeting clinical targets since enrollment (Figure 1B), especially for Ramsey. Compared with baseline, where fewer than 5% of patients with diabetes were meeting treatment targets, 25.6% (14.5-41.0%) of Ramsey HealthRise patients with diabetes had A1c levels lower than 8%. Yet many HealthRise patients with diabetes still had A1c levels of 8% or higher by endline across sites: 75.7% in Hennepin, 74.4% in Ramsey, and 49.0% in Rice. A1c category shifts between baseline and endline were especially varied for Hennepin and Rice; for nearly every A1c category at baseline (i.e., < 7%, 7-7.9%, 8-9.9%, ≥10%), some portion of patients moved to one of the other three A1c categories by endline.

Unadjusted and adjusted difference-in-difference model results were nearly identical for the effect of HealthRise (Table 4); accordingly, we report on the adjusted model results here. Overall, HealthRise patients trended toward greater progress in reducing biometric measures and meeting treatment targets than comparison patients; however, these differences were not consistently significant across indicators and sites. For hypertension patients, HealthRise participation was associated with statistically significant SBP reductions relative to comparison patients in Rice (6.9 mmHg decrease [0.9-13.0; *p* < 0.05]). Relative to comparison patients, HealthRise was also associated with a statistically significant increase in the percentage of hypertension patients meeting treatment targets in Hennepin (27.3 percentage-point rise [9.7-45.0; *p* < 0.01]) and Rice (17.1 percentage-point increase [0.9 to 33.4; *p* < 0.05]). In Ramsey, changes in hypertension indicators were not statistically different between HealthRise and comparison group, though program participation trended toward improvement: a 10.7 mmHg decrease (−0.3-21.8; *p*=0.057) in SBP and 22.3 percentage-point increase (−0.01 to 44.4=5; *p*=0.054) in meeting treatment targets since baseline.

**Table 4.**
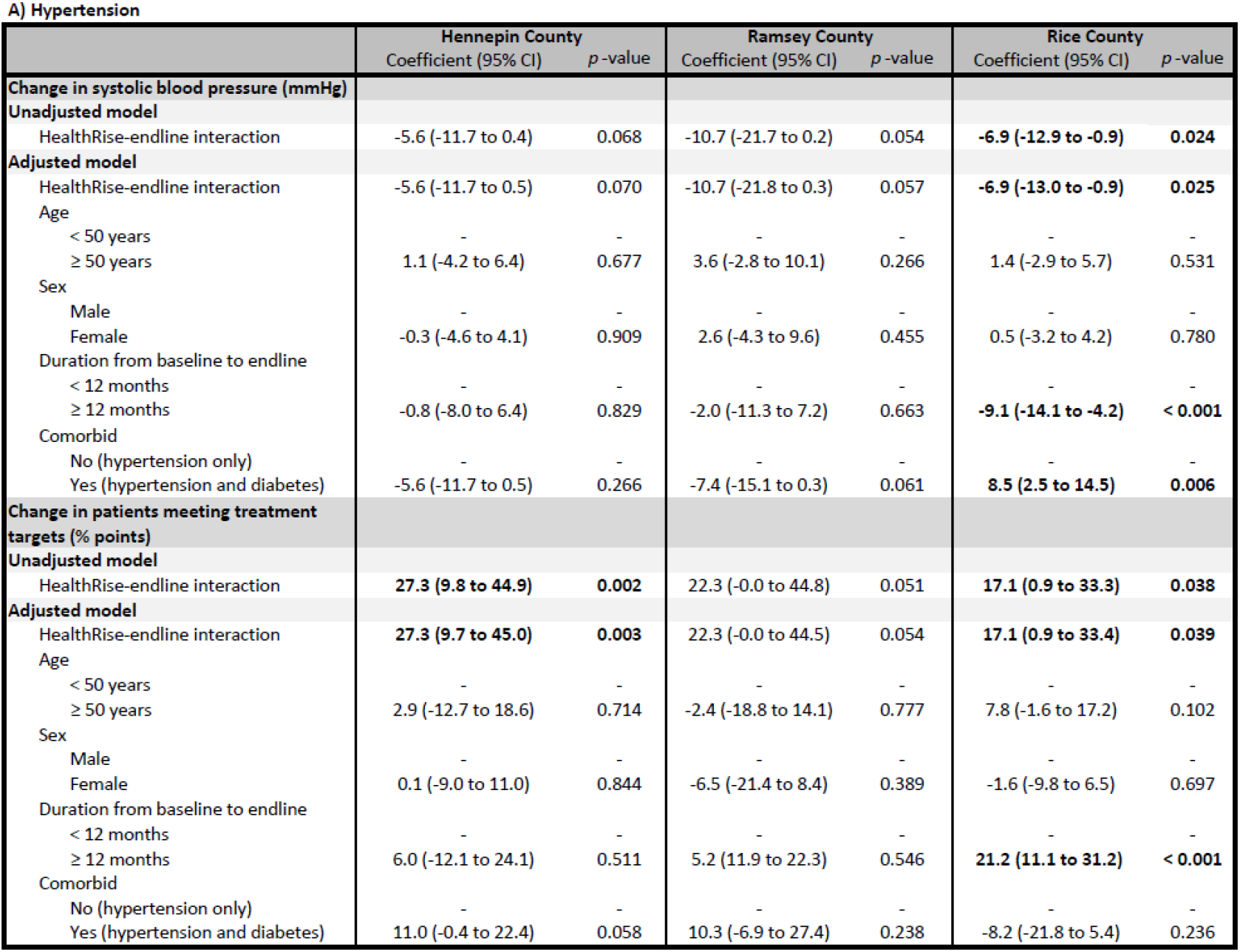
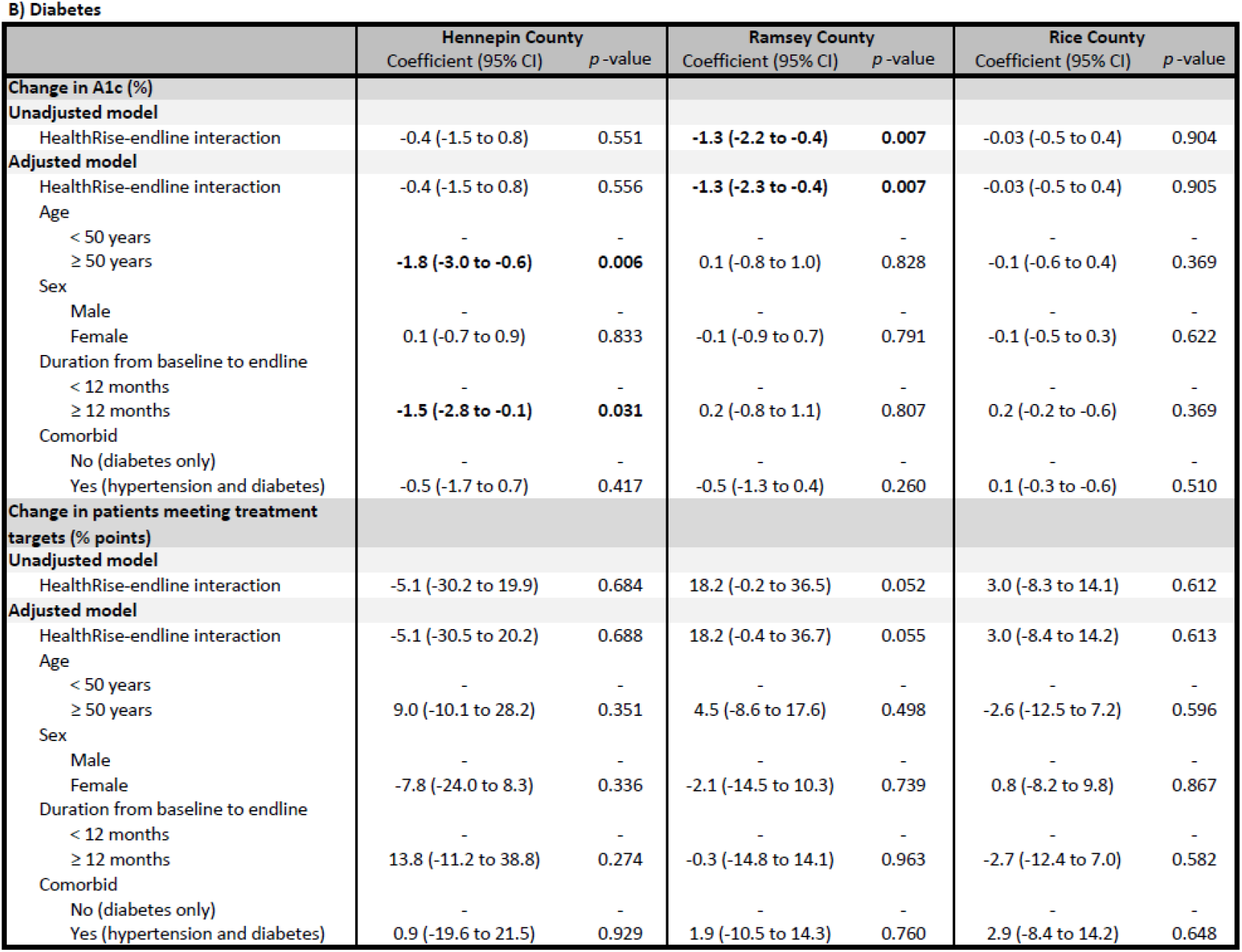
Unadjusted and adjusted difference-in-difference regression results, by HealthRise grantee, for hypertension (A) and diabetes (B) patients. Patient samples included in this analysis are those who met all inclusion critiera, remained enrolled throughout the program (for HealthRise patients), and had at least two biometric readings to reflect potential changes between baseline and endline. Bolded values reflect statistically significant estimates at *p* < 0.05.

**A) Hypertension**
|  | **Hennepin County** |  | **Ramsey County** |  | **Rice County** |  |
| --- | --- | --- | --- | --- | --- | --- |
|  | Coefficient (95% CI) | p-value | Coefficient (95% CI) | p-value | Coefficient (95% CI) | p-value |
| **Change in systolic blood pressure (mmHg)** |  |  |  |  |  |  |
| **Unadjusted model** |  |  |  |  |  |  |
| HealthRise-endline interaction | -5.6 (-11.7 to 0.4) | 0.068 | -10.7 (-21.7 to 0.2) | 0.054 | **-6.9 (-12.9 to -0.9)** | **0.024** |
| **Adjusted model** |  |  |  |  |  |  |
| HealthRise-endline interaction | -5.6 (-11.7 to 0.5) | 0.070 | -10.7 (-21.8 to 0.3) | 0.057 | **-6.9 (-13.0 to -0.9)** | **0.025** |
| Age |  |  |  |  |  |  |
| < 50 years | - | - | - | - | - | - |
| ≥ 50 years | 1.1 (-4.2 to 6.4) | 0.677 | 3.6 (-2.8 to 10.1) | 0.266 | 1.4 (-2.9 to 5.7) | 0.531 |
| Sex |  |  |  |  |  |  |
| Male | - | - | - | - | - | - |
| Female | -0.3 (-4.6 to 4.1) | 0.909 | 2.6 (-4.3 to 9.6) | 0.455 | 0.5 (-3.2 to 4.2) | 0.780 |
| Duration from baseline to endline |  |  |  |  |  |  |
| < 12 months | - | - | - | - | - | - |
| ≥ 12 months | -0.8 (-8.0 to 6.4) | 0.829 | -2.0 (-11.3 to 7.2) | 0.663 | **-9.1 (-14.1 to -4.2)** | **< 0.001** |
| Comorbid |  |  |  |  |  |  |
| No (hypertension only) | - | - | - | - | - | - |
| Yes (hypertension and diabetes) | -5.6 (-11.7 to 0.5) | 0.266 | -7.4 (-15.1 to 0.3) | 0.061 | **8.5 (2.5 to 14.5)** | **0.006** |
| **Change in patients meeting treatment targets (% points)** |  |  |  |  |  |  |
| **Unadjusted model** |  |  |  |  |  |  |
| HealthRise-endline interaction | **27.3 (9.8 to 44.9)** | **0.002** | 22.3 (-0.0 to 44.8) | 0.051 | **17.1 (0.9 to 33.3)** | **0.038** |
| **Adjusted model** |  |  |  |  |  |  |
| HealthRise-endline interaction | **27.3 (9.7 to 45.0)** | **0.003** | 22.3 (-0.0 to 44.5) | 0.054 | **17.1 (0.9 to 33.4)** | **0.039** |
| Age |  |  |  |  |  |  |
| < 50 years | - | - | - | - | - | - |
| ≥ 50 years | 2.9 (-12.7 to 18.6) | 0.714 | -2.4 (-18.8 to 14.1) | 0.777 | 7.8 (-1.6 to 17.2) | 0.102 |
| Sex |  |  |  |  |  |  |
| Male | - | - | - | - | - | - |
| Female | 0.1 (-9.0 to 11.0) | 0.844 | -6.5 (-21.4 to 8.4) | 0.389 | -1.6 (-9.8 to 6.5) | 0.697 |
| Duration from baseline to endline |  |  |  |  |  |  |
| < 12 months | - | - | - | - | - | - |
| ≥ 12 months | 6.0 (-12.1 to 24.1) | 0.511 | 5.2 (11.9 to 22.3) | 0.546 | **21.2 (11.1 to 31.2)** | **< 0.001** |
| Comorbid |  |  |  |  |  |  |
| No (hypertension only) | - | - | - | - | - | - |
| Yes (hypertension and diabetes) | 11.0 (-0.4 to 22.4) | 0.058 | 10.3 (-6.9 to 27.4) | 0.238 | -8.2 (-21.8 to 5.4) | 0.236 |

B) Diabetes
|  | Hennepin County |  | Ramsey County |  | Rice County |  |
| --- | --- | --- | --- | --- | --- | --- |
|  | Coefficient (95% CI) | p-value | Coefficient (95% CI) | p-value | Coefficient (95% CI) | p-value |
| **Change in A1c (%)** |  |  |  |  |  |  |
| **Unadjusted model** |  |  |  |  |  |  |
| HealthRise-endline interaction | -0.4 (-1.5 to 0.8) | 0.551 | **-1.3 (-2.2 to -0.4)** | **0.007** | -0.03 (-0.5 to 0.4) | 0.904 |
| **Adjusted model** |  |  |  |  |  |  |
| HealthRise-endline interaction | -0.4 (-1.5 to 0.8) | 0.556 | **-1.3 (-2.3 to -0.4)** | **0.007** | -0.03 (-0.5 to 0.4) | 0.905 |
| Age |  |  |  |  |  |  |
| < 50 years | - | - | - | - | - | - |
| ≥ 50 years | **-1.8 (-3.0 to -0.6)** | **0.006** | 0.1 (-0.8 to 1.0) | 0.828 | -0.1 (-0.6 to 0.4) | 0.369 |
| Sex |  |  |  |  |  |  |
| Male | - | - | - | - | - | - |
| Female | 0.1 (-0.7 to 0.9) | 0.833 | -0.1 (-0.9 to 0.7) | 0.791 | -0.1 (-0.5 to 0.3) | 0.622 |
| Duration from baseline to endline |  |  |  |  |  |  |
| < 12 months | - | - | - | - | - | - |
| ≥ 12 months | **-1.5 (-2.8 to -0.1)** | **0.031** | 0.2 (-0.8 to 1.1) | 0.807 | 0.2 (-0.2 to -0.6) | 0.369 |
| Comorbid |  |  |  |  |  |  |
| No (diabetes only) | - | - | - | - | - | - |
| Yes (hypertension and diabetes) | -0.5 (-1.7 to 0.7) | 0.417 | -0.5 (-1.3 to 0.4) | 0.260 | 0.1 (-0.3 to -0.6) | 0.510 |
| **Change in patients meeting treatment targets (% points)** |  |  |  |  |  |  |
| **Unadjusted model** |  |  |  |  |  |  |
| HealthRise-endline interaction | -5.1 (-30.2 to 19.9) | 0.684 | 18.2 (-0.2 to 36.5) | 0.052 | 3.0 (-8.3 to 14.1) | 0.612 |
| **Adjusted model** |  |  |  |  |  |  |
| HealthRise-endline interaction | -5.1 (-30.5 to 20.2) | 0.688 | 18.2 (-0.4 to 36.7) | 0.055 | 3.0 (-8.4 to 14.2) | 0.613 |
| Age |  |  |  |  |  |  |
| < 50 years | - | - | - | - | - | - |
| ≥ 50 years | 9.0 (-10.1 to 28.2) | 0.351 | 4.5 (-8.6 to 17.6) | 0.498 | -2.6 (-12.5 to 7.2) | 0.596 |
| Sex |  |  |  |  |  |  |
| Male | - | - | - | - | - | - |
| Female | -7.8 (-24.0 to 8.3) | 0.336 | -2.1 (-14.5 to 10.3) | 0.739 | 0.8 (-8.2 to 9.8) | 0.867 |
| Duration from baseline to endline |  |  |  |  |  |  |
| < 12 months | - | - | - | - | - | - |
| ≥ 12 months | 13.8 (-11.2 to 38.8) | 0.274 | -0.3 (-14.8 to 14.1) | 0.963 | -2.7 (-12.4 to 7.0) | 0.582 |
| Comorbid |  |  |  |  |  |  |
| No (diabetes only) | - | - | - | - | - | - |
| Yes (hypertension and diabetes) | 0.9 (-19.6 to 21.5) | 0.929 | 1.9 (-10.5 to 14.3) | 0.760 | 2.9 (-8.4 to 14.2) | 0.648 |

Among diabetes patients, HealthRise participation was associated with statistically significant reductions in A1c in Ramsey (1.3 decrease in A1c [0.4-3.2; *p* < 0.01) relative to comparison patients. While the percentage of HealthRise patients meeting treatment targets for diabetes did not statistically differ from that of comparison patients in Ramsey, this indicator trended toward improvement as well (a 18.2 percentage-point increase [-0.4-36.7; *p*=0.054]).

### Qualitative findings

Across HealthRise sites, six main themes emerged for the qualitative data synthesis (Table 5). First, respondents viewed home-based providers as critical to bridging barriers experienced by patients (e.g., linguistic and cultural divides), and clinic-based providers indicated high value in meeting patients beyond clinical settings. Coordination of care and a focus on social determinants of health, such as access to healthier food and nutrition, were highlighted as key program features. Second, program strengths involved learning from HealthRise sites in other countries and enabling many clinical staff to work with in-home providers for the first time. Clinical providers reported improved quality and efficiency in clinical appointments due to having additional details about patient needs from in-home providers. Home visits also enabled providers to connect patients with non-clinical resources (e.g., housing) to support improved outcomes. The theme of perceived program impacts extended program strengths, with providers reporting positive changes in the health and lives of patients and their families. Further, several interviewees emphasized the synergistic effects of pairing CHWs and community paramedics (CPs) within care teams, and reported efforts to adopt home-based provider models by other local organizations because of HealthRise experiences.

**Table 5.**
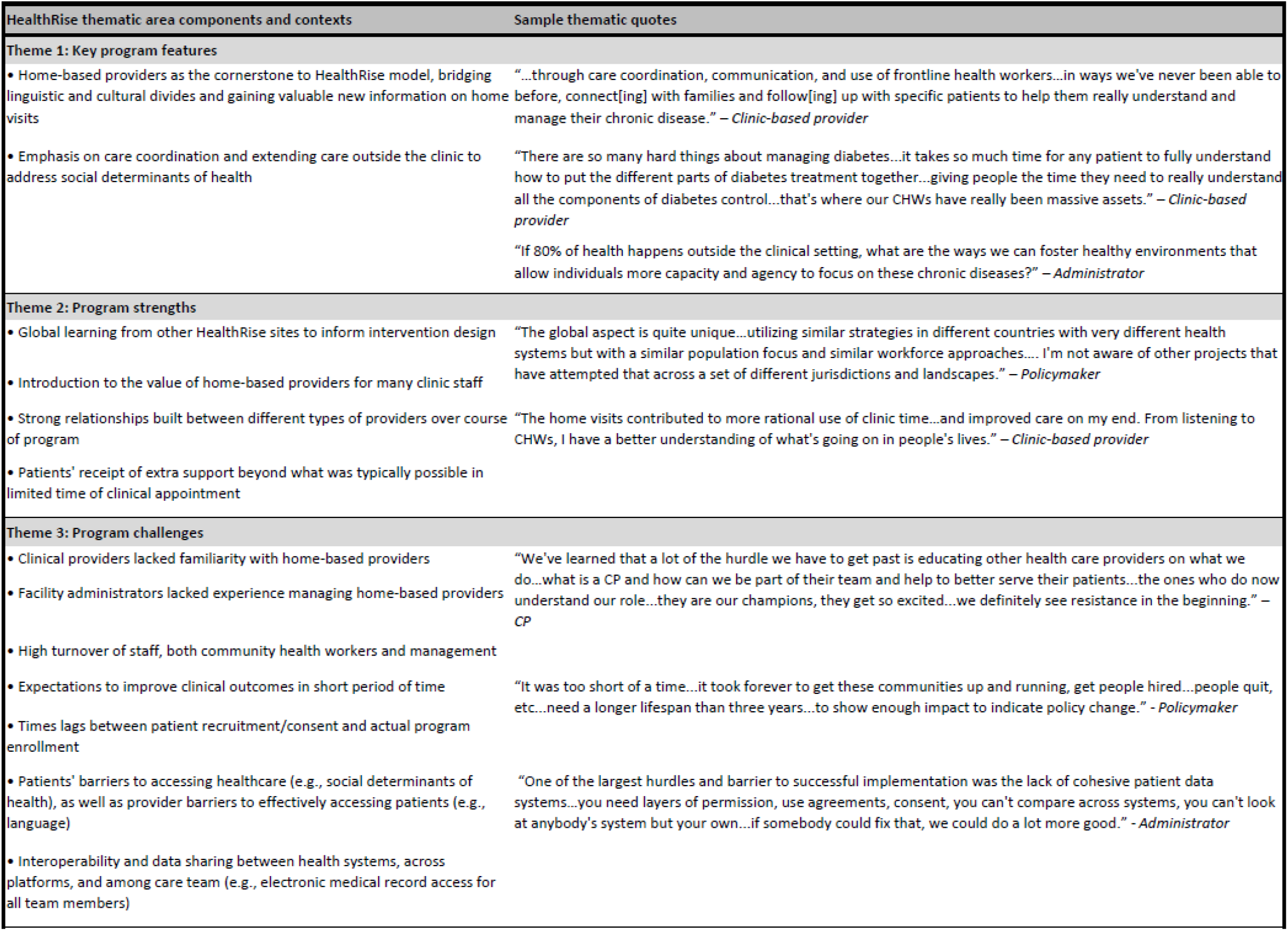
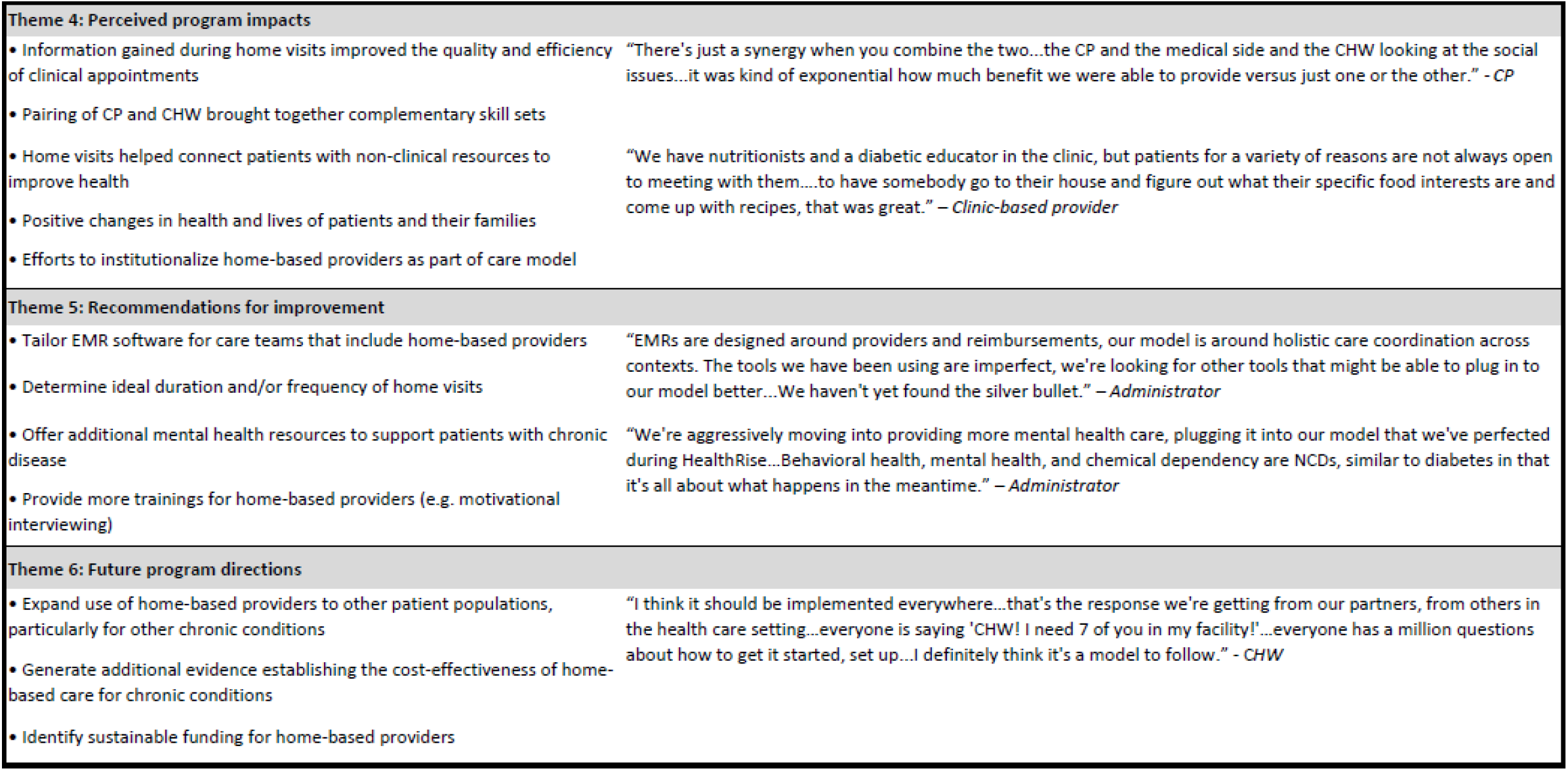
Summary of key themes, components, and quotes from qualitative data synthesized across US HealthRise sites.

| HealthRise thematic area components and contexts | Sample thematic quotes |
| --- | --- |
| **Theme 1: Key program features** |  |
| • Home-based providers as the cornerstone to HealthRise model, bridging linguistic and cultural divides and gaining valuable new information on home visits<br>• Emphasis on care coordination and extending care outside the clinic to address social determinants of health | "...through care coordination, communication, and use of frontline health workers...in ways we've never been able to before, connect[ing] with families and follow[ing] up with specific patients to help them really understand and manage their chronic disease." – *Clinic-based provider*<br>"There are so many hard things about managing diabetes...it takes so much time for any patient to fully understand how to put the different parts of diabetes treatment together...giving people the time they need to really understand all the components of diabetes control...that's where our CHWs have really been massive assets." – *Clinic-based provider*<br>"If 80% of health happens outside the clinical setting, what are the ways we can foster healthy environments that allow individuals more capacity and agency to focus on these chronic diseases?" – *Administrator* |
| **Theme 2: Program strengths** |  |
| • Global learning from other HealthRise sites to inform intervention design<br>• Introduction to the value of home-based providers for many clinic staff<br>• Strong relationships built between different types of providers over course of program<br>• Patients' receipt of extra support beyond what was typically possible in limited time of clinical appointment | "The global aspect is quite unique...utilizing similar strategies in different countries with very different health systems but with a similar population focus and similar workforce approaches.... I'm not aware of other projects that have attempted that across a set of different jurisdictions and landscapes." – *Policymaker*<br>"The home visits contributed to more rational use of clinic time...and improved care on my end. From listening to CHWs, I have a better understanding of what's going on in people's lives." – *Clinic-based provider* |
| **Theme 3: Program challenges** |  |
| • Clinical providers lacked familiarity with home-based providers<br>• Facility administrators lacked experience managing home-based providers<br>• High turnover of staff, both community health workers and management<br>• Expectations to improve clinical outcomes in short period of time<br>• Times lags between patient recruitment/consent and actual program enrollment<br>• Patients' barriers to accessing healthcare (e.g., social determinants of health), as well as provider barriers to effectively accessing patients (e.g., language)<br>• Interoperability and data sharing between health systems, across platforms, and among care team (e.g., electronic medical record access for all team members) | "We've learned that a lot of the hurdle we have to get past is educating other health care providers on what we do...what is a CP and how can we be part of their team and help to better serve their patients...the ones who do now understand our role...they are our champions, they get so excited...we definitely see resistance in the beginning." – *CP*<br>"It was too short of a time...it took forever to get these communities up and running, get people hired...people quit, etc...need a longer lifespan than three years...to show enough impact to indicate policy change." – *Policymaker*<br>"One of the largest hurdles and barrier to successful implementation was the lack of cohesive patient data systems...you need layers of permission, use agreements, consent, you can't compare across systems, you can't look at anybody's system but your own...if somebody could fix that, we could do a lot more good." – *Administrator* |

| HealthRise thematic area components and contexts (*continued*) | Sample thematic quotes |
| --- | --- |
| **Theme 4: Perceived program impacts** |  |
| Information gained during home visits improved the quality and efficiency of clinical appointments<br>Pairing of CP and CHW brought together complementary skill sets<br>Home visits helped connect patients with non-clinical resources to improve health<br>Positive changes in health and lives of patients and their families<br>Efforts to institutionalize home-based providers as part of care model | "There's just a synergy when you combine the two...the CP and the medical side and the CHW looking at the social issues...it was kind of exponential how much benefit we were able to provide versus just one or the other." - CP<br>"We have nutritionists and a diabetic educator in the clinic, but patients for a variety of reasons are not always open to meeting with them....to have somebody go to their house and figure out what their specific food interests are and come up with recipes, that was great." – *Clinic-based provider* |
| **Theme 5: Recommendations for improvement** |  |
| Tailor EMR software for care teams that include home-based providers<br>Determine ideal duration and/or frequency of home visits<br>Offer additional mental health resources to support patients with chronic disease<br>Provide more trainings for home-based providers (e.g. motivational interviewing) | "EMRs are designed around providers and reimbursements, our model is around holistic care coordination across contexts. The tools we have been using are imperfect, we're looking for other tools that might be able to plug in to our model better...We haven't yet found the silver bullet." – *Administrator*<br>"We're aggressively moving into providing more mental health care, plugging it into our model that we've perfected during HealthRise...Behavioral health, mental health, and chemical dependency are NCDs, similar to diabetes in that it's all about what happens in the meantime." – *Administrator* |
| **Theme 6: Future program directions** |  |
| Expand use of home-based providers to other patient populations, particularly for other chronic conditions<br>Generate additional evidence establishing the cost-effectiveness of home-based care for chronic conditions<br>Identify sustainable funding for home-based providers | "I think it should be implemented everywhere...that's the response we're getting from our partners, from others in the health care setting...everyone is saying 'CHW! I need 7 of you in my facility!'...everyone has a million questions about how to get it started, set up...I definitely think it's a model to follow." - CHW |

Common challenges emerged across sites, often relating to new program establishment and incorporation of in-home providers within care teams. For example, some clinical providers showed initial skepticism about the added value of in-home providers, and most administrators did not have prior experience managing CHWs and CPs. Site-specific challenges also occurred; for instance, patient consent for enrollment took a long time during the initial phase of program implementation in Rice, while CHW turnover was an ongoing obstacle for both Hennepin and Ramsey. Data sharing was another pervasive challenge, mainly from poor interoperability and coordination between clinic data systems and electronic medical record (EMR) systems. Providers also expressed frustration with expectations for rapidly improving clinical outcomes, especially given the longstanding challenges in healthcare access and social determinants of health most patients faced.

The fifth theme pertained to recommendations for improvement, many of which stemmed from acknowledged challenges. Such suggestions included prioritizing better communication and coordination among care teams as well as EMR systems that could more seamlessly accommodate patient updates and provider notes from multiple care-team members. Interviewees reported having inadequate clarity on the ideal frequency and length of home visits, an area where efficiencies in resource deployment could be improved. The sixth theme, future program directions, involved many ideas about adapting and expanding HealthRise programming to new locations and conditions (e.g., mental health). Another common thread concerned program sustainability, namely longer-term financing and retaining home-based providers for the HealthRise model.

## Discussion

Increasingly more evidence shows that community-based programs can help underserved communities in the US better access health services, alleviate barriers to care, and improve at least some health behaviors and outcomes. The present study contributes to this evidence base through its prospective evaluation of HealthRise programs in three Minnesota communities. Relative to comparison patients, HealthRise participation was significantly associated with SBP reductions in Rice, while the percentage of hypertension patients meeting treatment targets increased in Hennepin and Rice. For diabetes, HealthRise patients saw larger A1c declines in Ramsey than comparison patients. Heterogeneous patterns in patient improvements since baseline highlight potential case management challenges among underserved individuals and communities, especially under short program implementation periods. As emphasized by HealthRise care teams, community-based programs show promise for improving NCD care and outcomes for underserved populations; nonetheless, more work is needed to better understand how such programs can be further brought to scale and sustained long-term.

HealthRise participation was related to SBP or A1c decreases and a higher percentage of patients meeting treatment targets at some sites relative to comparison patients. Variations across sites may be related to different demographic characteristics or relative health status of each population at baseline. For instance, Ramsey HealthRise patients with diabetes averaged 11.4% reductions in A1c and 9.4% declines in SBP by endline; yet Ramsey patients also began HealthRise with highest risk profiles, averaging 150 mmHg SBP and 10% A1c at baseline. As found in past studies,[5–7] several factors may have contributed to the observable effects of HealthRise. These included focusing on specific barriers patients faced in each community (e.g., home-based care provided by CHW and CP teams); maintaining small patient loads, enabling more individualized attention and tailoring of visit frequency to patient need; and explicitly providing non-medical support, such as health education and community resources like transportation. Further, the overall positive views of HealthRise by grantees and care teams alike may have contributed to the program’s effects. Despite challenges during earlier stages of program implementation (e.g., recruiting and retaining CHWs, ensuring adequate access to EMRs), providers voiced valuing home-based health workers and were eager to expand this model of care. In combination, these factors may have set the foundation for HealthRise’s impact for underserved patients with hypertension and diabetes in the US.

Amid such promising findings, however, important challenges remained for each site and for broader applications of the HealthRise model elsewhere. Across sites, some proportion of HealthRise patients failed to meet clinical targets for hypertension or diabetes at both time points – and concerningly, some percentage moved from being below biometric thresholds at baseline to exceeding them at endline. These patterns underscore the complexity of effectively managing chronic conditions like hypertension and diabetes, especially in environments where patients face compounding obstacles to medical care and health-promoting behavior (e.g., limited budgets for nutritious food, minimal time for exercise amid job and family demands, inferior access to adequate transportation and housing). CHWs and integrated care teams may be able to mitigate some of these barriers and better support patients’ medical needs, a critical step in addressing deep-seeded health disparities; however, in the absence of more macro-level socioeconomic policies and health system investments to support underserved patients in the US, many community-based programs will continue facing need and demand that far exceeds their limited capacities and resources. As laid bare by COVID-19, the health challenges underserved communities experience do not begin and end at clinics: rather, they stem from and are exacerbated by structural inequalities that require intervention and engagement well beyond the formal health system.[20,21] CHWs can provide a vital role in at least overcoming some of these access and sociocultural barriers, ranging from house-based visits made by CHWs fluent in patients’ native languages[3] to connecting patients with services that can facilitate better overall care. Nevertheless, the potential benefits of encouraging health programs can be easily blunted if actors – and actions – beyond the immediate health system are not also actively addressing fundamental drivers of health disparities.

### Limitations

This study is subject to several limitations. First, small study samples at each site likely affected the degree to which potential program impact could be detected and conclusively attributed to HealthRise participation. This is particularly true for site-condition combinations in which very few patients were prevalent cases at baseline and had at least two biometric readings within the program implementation period (e.g., 37 HealthRise patients with diabetes in Hennepin, 32 HealthRise patients with hypertension in Ramsey). The inclusion or exclusion of even a few patients for several site-condition groupings could shift effect sizes and statistical significance estimated by the difference-in-difference models.

Second, comparison groups were constructed retrospectively based on available patient record information and were not selected by random assignment. While efforts were made to ensure that comparison patient data were chosen to generally represent individuals who would have been eligible for HealthRise enrollment, they may have differed from individuals who enrolled.

Third, only patients who remained enrolled at endline were included in the present study; by taking this ‘as treated’ analytic approach, which provides insights into program effects closer to full adherence, these patients may not represent all potential target populations for HealthRise interventions and results may be positively biased. For instance, relatively high rates of program withdrawal at some sites could have led to a bias toward healthier patients remaining in the program (i.e., sicker patients may not go into the clinic). However, due to the home-based care model espoused by HealthRise, it is equally possible that patients who did not withdraw were less healthy and stayed enrolled because HealthRise offered important access to services, like home visits, otherwise unavailable to them.

Fourth, single biometric readings comprised baseline and endline measures, as well as patients meeting clinical targets at each time period; subsequently, analyses could be sensitive to outliers in patient records, particularly given the relatively small sample sizes for each site. If more readings could have informed baseline and endline indicators, it is possible patient-level patterns could have differed from what observed on the basis of single readings.

Fifth, medication data were not available and thus we were unable to assess the full cascade of care for hypertension and diabetes case management. Medication adherence may have been important factor for patients who either failed to see improvements in clinical indicators or experienced worsening outcomes.

Sixth, due to the small sample sizes for each site, we could not further analyze the potential effects of visit frequency (and thus approximate dose-response) or intensity of intervention exposure on patient outcomes. This is further complicated by issues related to endogeneity, such that patients with worse clinical profiles and thus greater need are likely to receive more frequent visits by CHWs or care teams.

Seventh, small sample sizes also limited our ability to conduct more disaggregated analyses by sex, race or ethnicity, income, and other important characteristics for understanding how community-based health interventions can promote greater equity for underserved communities.

### Conclusions

With its focus on community-based health programs and improving NCD care for underserved populations, HealthRise showed some positive effects for hypertension and diabetes patients in Hennepin, Ramsey, and Rice counties. Provider experiences indicated enthusiasm for expanding the home-based model of care for NCDs, though the resource requirements – as well feasibility – to sustain impact at a larger scale remain unknown. While community-based NCD interventions show promise for overcoming barriers to effective care for hypertension and diabetes among underserved populations, continued monitoring and robust evaluations of local impact are vital to ensuring maximum benefit for individuals with the greatest need.

## Data Availability

Data are available in a public, open access repository. The datasets generated and/or
analysed during the current study are available, when possible, in the GHDx data
repository, http://ghdx.healthdata.org/series/healthrise-evaluation

## Acknowledgments

We would like to acknowledge the support and insights from the US HealthRise program implementers at Pillsbury United Communities, Regions Hospital Foundation, and HealthFinders Collaborative Inc., of whom also provided the de-identified data for this study. Without them, this work would not be possible. We also acknowledge the state and local governments in HealthRise areas for their cooperation and support with HealthRise intervention and evaluation activities. We thank Kelsey Bannon, Erin Palmisano, and Laura Di Giorgio for assisting in the management and execution of the HealthRise evaluation. Last, we thank all individuals who participated in this study.

